# Novel Ultra-Rare Exonic Variants Identified in a Founder Population Implicate Cadherins in Schizophrenia

**DOI:** 10.1101/2020.05.29.20115352

**Authors:** Todd Lencz, Jin Yu, Raiyan Rashid Khan, Shai Carmi, Max Lam, Danny Ben-Avraham, Nir Barzilai, Susan Bressman, Ariel Darvasi, Judy H. Cho, Lorraine N. Clark, Zeynep H. Gümüş, Joseph Vijai, Robert J. Klein, Steven Lipkin, Kenneth Offit, Harry Ostrer, Laurie J. Ozelius, Inga Peter, Anil K. Malhotra, Gil Atzmon, Itsik Pe’er

## Abstract

Identification of rare genetic variants associated with schizophrenia has proven challenging due to multiple sources of heterogeneity, which may be reduced in founder populations. We examined ultra-rare exonic variants in 786 patients with schizophrenia and 463 healthy comparison subjects, all drawn from the Ashkenazi Jewish population. Cases had a higher frequency of novel missense or loss of function (MisLoF) variants compared to controls. Characterizing 141 “case-only” genes (in which ≥ 3 cases in our dataset had MisLoF variants with none found in controls), we identified cadherins as a novel gene set associated with schizophrenia, including a recurrent mutation in *PCDHA3*. Modeling the effects of purifying selection demonstrated that deleterious ultra-rare variants are greatly over-represented in the Ashkenazi population, resulting in enhanced power for rare variant association. Identification of cell adhesion genes in the cadherin/protocadherin family helps specify the synaptic abnormalities central to the disorder, and suggests novel potential treatment strategies.

---

Twin studies and other family-based designs have long demonstrated that schizophrenia (SCZ) is highly heritable (h^2^≈.6-.85) (Hilker et al., 2018; McGue et al., 1983; Sullivan et al., 2003). While large-scale genome-wide association studies (GWAS) have discovered increasing numbers of common (minor allele frequency > 1%) variants associated with illness (Lam et al., 2019; Pardiñas et al., 2018; Schizophrenia Working Group of the Psychiatric Genomics Consortium, 2014), the cumulative effect of such variants accounts for only about a third of the total heritability of SCZ (Lee et al., 2012; Loh et al., 2015). It is therefore likely that rare genetic variants contribute substantially to the heritability of SCZ (Ganna et al., 2018; Purcell et al., 2014), and such rare variants might have considerably higher effect sizes (odds ratios) relative to common variants (Sullivan et al., 2012). For example, several rare (frequency<<1% in the general population) copy number variants have been reliably associated with SCZ, with odds ratios ranging from 5-20 or higher (Marshall et al., 2017).

Identification of rare single nucleotide variants (SNVs) associated with SCZ has proven difficult for several reasons: 1) SCZ is marked by a high degree of locus heterogeneity due to the large “mutational target” (i.e., damage to many different genes can increase risk for the phenotype) (Gratten et al., 2014); 2) at any given gene, a variety of different alleles may have deleterious effects (allelic heterogeneity) (Li & Leal, 2009); 3) deleterious rare variants are generally driven to extremely low frequencies due to purifying selection (Kryukov et al., 2007); and 4) the background rate of benign rare variation across the population is very high (Tennessen et al., 2012). To date, only very large international consortia efforts have identified any schizophrenia-associated SNV’s. The largest such effort, the Schizophrenia Exome Sequencing Meta-analysis (SCHEMA) consortium with 25,000 cases and nearly 100,000 controls, identified only 10 exome-wide significant genes (Kaiser et al., 2019).

One approach to enhance power in rare variant studies is to examine unusual populations marked by a strong, (relatively) recent founder effect; such populations are enriched for deleterious rare variants due to inefficient purifying selection (Locke et al., 2019; Wang et al., 2014). For example, the Ashkenazi Jewish (AJ) population, currently numbering more than 10 million individuals worldwide, effectively derives from a mere ~300 founders approximately 750 years ago (Carmi et al., 2014; Palamara et al., 2012). While the AJ population is well known to be enriched for deleterious variants leading to rare recessive disorders (Baskovich et al., 2016), AJ also demonstrate a 10-fold elevated frequency of high-penetrance risk variants for common complex disease, such as the *LRRK2* p.G2019S allele associated with Parkinson’s disease (Ozelius et al., 2006) and the *BRCA1* c.66_67AG allele associated with breast cancer (Friedman et al., 1995). Importantly, a recent large-scale (n>5,000) sequencing study of AJ individuals demonstrated that this enrichment is widespread across the exome, with approximately one-third of all protein-coding alleles demonstrating frequencies in AJ that were an order of magnitude greater than the maximum frequency in any well-characterized outbred population (Rivas et al., 2018).

In the present study, we examined rates of protein-altering rare variants in AJ cases with schizophrenia compared with AJ controls. Based on prior research (Genovese et al., 2016; Gulsuner et al., 2020; Nguyen et al., 2017; Purcell et al., 2014), we hypothesized that cases would be enriched for rare deleterious variants, especially in genes expressed at the neuronal synapse. We sought to extend these results to additional categories of genes that might be detectable due to the greater frequency of rare variants observed in the AJ population. Additionally, we attempted to replicate the schizophrenia risk genes identified by the SCHEMA consortium. Finally, we modelled the process of purifying selection in a rapidly expanding, bottlenecked population, in order to quantify the relative power of AJ for rare variant discovery.

## Results

### Greater Rates of Damaging Variants in Cases

After QC procedures, a total of 786 SCZ cases and 463 controls were available for final analysis. Groups did not significantly differ in total number of variants called in their whole genome (~3.68M) or exome (~49K) (Table 1). After filtering on all variants observed in theTOPMED and gnomAD (v2.1.1, non-neuro) datasets, cases and controls did not significantly differ on total number of novel variants observed genome-wide (~5K). However, cases had significantly more novel exome-wide variants, exclusively limited to singletons (17.76±6.24 vs 15.44±6.42, p=6.13×10^−10^; Table 1).

**Table 1.**

| **TABLE 1** |  | **per SCZ**<br>**case**<br>**(n=786)** | **s.d.** | **per**<br>**Control**<br>**(n=463)** | **s.d.** | **p-value** | **95% CI of**<br>**differences**<br>**(case - control)** |
| --- | --- | --- | --- | --- | --- | --- | --- |
| Post-QC | variants per genome | 3676280 | 17055 | 3676451 | 18899 | 0.87 | [-2268, 1925] |
|  | variants per exome | 49437 | 356 | 49454 | 387 | 0.45 | [-60, 27] |
| Post-Filter | variants per genome | 5028 | 427 | 5034 | 322 | 0.81 | [-47, 37] |
| | variants per exome | 30.64 | 6.14 | 28.66 | 7.15 | $1.55 \times 10^{-6}$ | [1.18, 2.79] |
|  | non-singleton variants per exome | 12.88 | 2.70 | 13.23 | 3.01 | 0.043 | [-0.68, -0.012] |
| | singleton variants (URV) per exome | 17.76 | 6.24 | 15.44 | 6.42 | $6.13 \times 10^{-10}$ | [1.59, 3.05] |
| | non-MisLoF | 9.86 | 4.13 | 8.81 | 3.97 | $8.42 \times 10^{-6}$ | [0.59, 1.52] |
| | MisLoF | 7.90 | 3.54 | 6.63 | 3.51 | $1.01 \times 10^{-9}$ | [0.87, 1.68] |

Against this backdrop, cases and controls were compared on the number of functional vs silent variants within the exome, in two ways: variant-based tests and gene-based tests. First, at the variant level, cases manifest a significantly elevated rate of novel MisLoF URVs (Table 1, last row). Cases also demonstrated an elevated rate of novel non-MisLoF URVs (Table 1, second-to-last row); however, even compared to this background elevation of non-MisLoF URVs, there was a significantly elevated proportion of exonic variants classified as MisLoF in cases (Fisher’s exact p=0.034). Next, we examined MisLoF and non-MisLoF URVs at the gene level. As shown in Figure 1, this ratio was much greater (i.e., more case-only genes than control-only genes) for MisLoF URVs relative to non-MisLoF URVs (p=0.0001 by permutation test).

**Figure 1.**
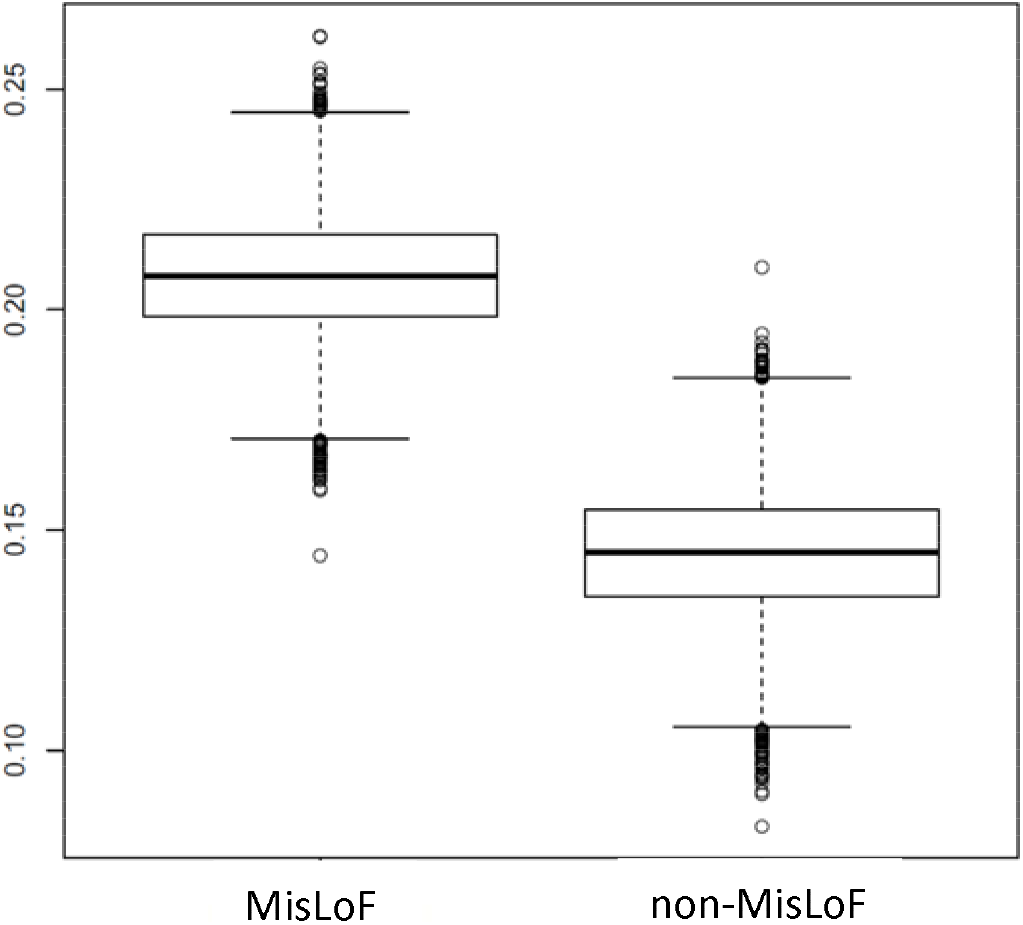
More Genes are Hit by Damaging URVs in Cases Relative to Controls. Y-axis denotes the degree of elevation of “case-only” genes to “control-only” genes. While there are more “case-only” genes hit by non-MisLoF variants, the case-control difference is much more pronounced for MisLoF genes (p=0.0001). Note that, for this analysis, we down-sampled the number of cases to match the number of controls, and iterated across 10,000 permutations.

### Replication of Previously Identified Schizophrenia Risk Genes

Given that cases demonstrated significant elevation in genes carrying MisLoF URVs, we next sought to characterize the case-only MisLoF genes. As shown in Supplementary Table 1, eight genes had Case ≥ 5 and Control = 0. Notably, one of these was *SETD1A*, a methyltransferase gene that was the first to reach genome-wide significance in a schizophrenia rare variant study (Singh et al., 2016). We next tested the set of 9 autosomal exome-wide-significant schizophrenia genes identified by the SCHEMA consortium (SCHEMA Consortium, 2020) for overlap with 141 “case-only” genes in which ≥ 3 AJ cases in our dataset had MisLoF URVs (with none found in our AJ controls). Three of 9 autosomal exome-wide significant SCHEMA genes (*SETD1A, TRIO*, and *XPO7*) were among the 141 case-only genes, a 47-fold over-representation relative to chance (hypergeometric test p = 3.03×10^−7^). Results remained significant when permutation tests controlling for gene size were performed (empirical p=5.8×10^−3^). Similar results were obtained examining overlap of our 141 case-only genes with the set of 29 autosomal SCHEMA genes that met the criteria of FDR<.05; in addition to the three genes above, *STAG1* was also shared between our case-only list and SCHEMA (4/29 genes; hypergeometric p=1.77×10^−6^; empirical p=4.6×10^−3^ using permutations controlling for gene size).

### Case-only Genes are Enriched for Known Neurodevelopmental Genes, Synaptic Genes, and Cadherins

More broadly, we utilized gene set analyses to characterize our 141 case-only genes, as compared to a similarly-sized set of 148 “control-only” genes in which ≥ 2 AJ controls had MisLoF URVs (with none found in our AJ cases). First, we tested sets of genes selected *a priori* based on prior literature; specifically, previous case-control exome studies in schizophrenia have identified: 1) overlaps with other developmental brain disorders (DBD) including autism spectrum disorder (ASD) and intellectual disability (ID); 2) gene sets representing critical synaptic and/or neurodevelopmental functions such as binding partners of FRMP, RBFOX, and CELF4; and 3) constrained genes (i.e., genes with far fewer missense and/or loss of function variants than average, presumably due to purifying selection) (Genovese et al., 2016; Gonzalez-Mantilla et al., 2016; Gulsuner et al., 2020; Nguyen et al., 2017; Purcell et al., 2014). As shown in Table 2a, each of these gene sets demonstrated significant (by hypergeometric test) overlap with the case-only, but not the control-only, gene lists; moreover, the difference in enrichment between the case-only and control-only gene sets was statistically significant in all cases. Permutation tests accounting for gene size demonstrated similar results, with the exception that the overlap with ASD/ID gene sets was no longer significant (Supplementary Table 2).

Next, we examined enrichment of our case-only gene list across all GO categories, Panther protein classes, and synaptic components (annotated by SYNGO (Koopmans et al., 2019)). As shown in Table 2b, novel categories of enrichment for the 141 schizophrenia-associated genes were observed for biological processes related to cell adhesion, and specifically to the cadherin class of proteins. GO cellular components analysis revealed the expected enrichment for synaptic and related compartments such as neuron projection and vesicle (Supplementary Table 3), while SYNGO analysis demonstrated that enrichment extended across both presynaptic and postsynaptic genes (Supplementary Table 4). By contrast, the 148 control-only genes showed no synaptic enrichment (all q-value>.75), no enriched GO categories (all FDR>.05), and only one enriched Panther protein class, which was in a very small gene set (Hsp90 family chaperone, overlap of 3/9 genes; p=3.59×10^−5^, FDR=7.00×10^−3^).

**Table 2.**
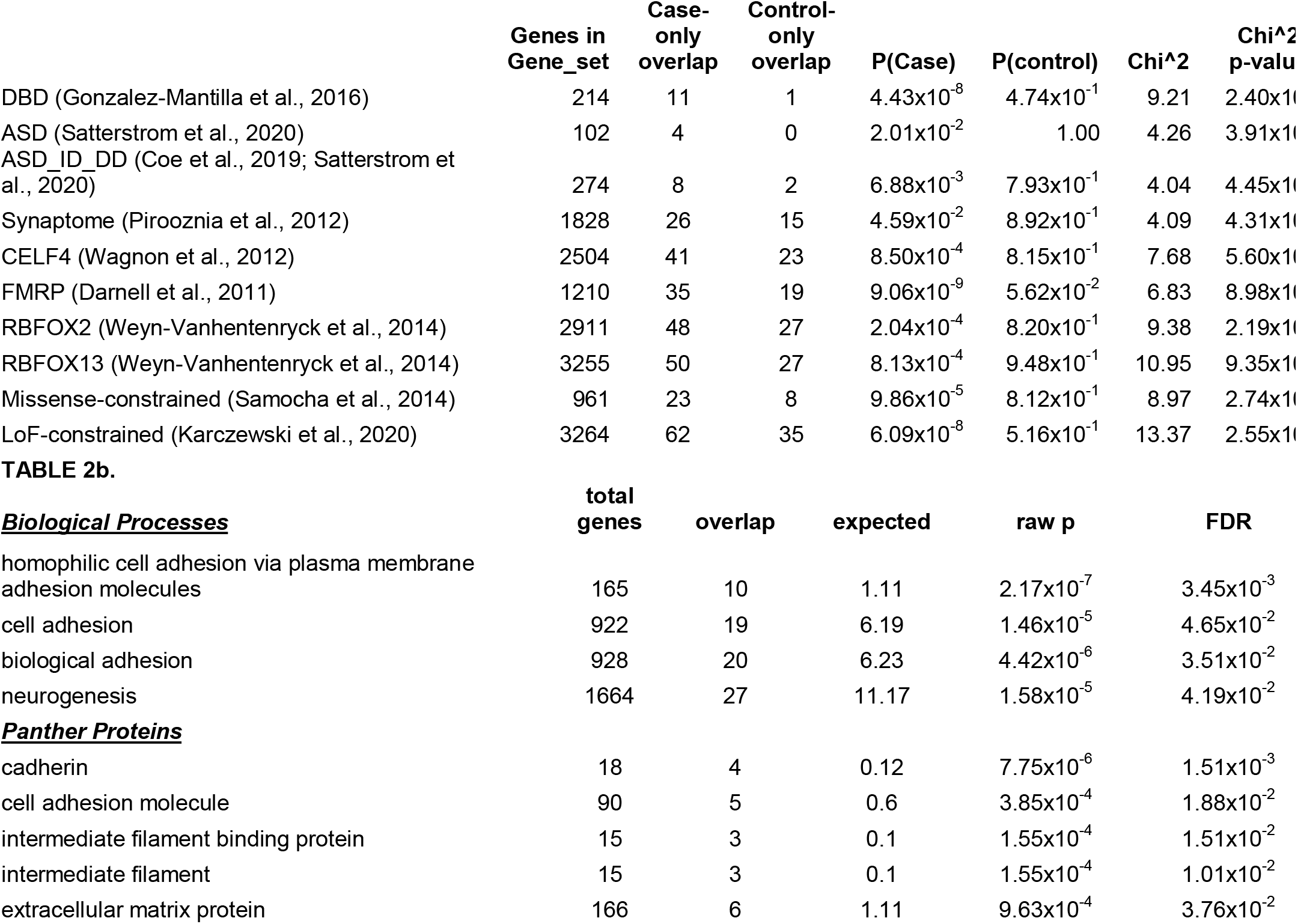

### A Damaging URV in PCDHA3 Is Observed Recurrently in Ashkenazi Schizophrenia Cases

The foregoing analyses examined singleton URVs only, which have been the primary focus of exome studies in schizophrenia to date; indeed, the SCHEMA dataset of case-only variants contains ~95% singletons, and <1% of all case-only variants in SCHEMA are observed 3 or more times. However, we hypothesized that the AJ population would be more likely than non-founder populations to retain and propagate multiple copies of deleterious variants. Consequently, we merged exome data from our cohort with the AJ schizophrenia cases (n=869) and controls (n=2415) from SCHEMA, in order to identify individual URVs that were observed ≥3 times in cases (i.e., at least ~1/1000 allele frequency in the 3,310 AJ case chromosomes available). For further filtering, we included exome data from an additional 1,587 AJ controls from a separate study of longevity, resulting in 8930 AJ control chromosomes available. As shown in Supplementary Table 6,17 MisLoF URVs were observed in ≥3 cases and zero controls (nominal p<.05 by Fisher’s exact test). Notably, the most common variant (observed 5 times, case frequency =.15%) is a putatively damaging (CADD score = 23.4) missense variant in *PCDHA3*, part of the protocadherin cluster on chromosome 5.

### Damaging Rare Variants Escape Purifying Selection in a Founder Population

Given that we were able to detect numerous recurrent case-only variants, despite our relatively small sample size compared to SCHEMA, we sought to model the parameters affecting the persistence of deleterious alleles in a founder population. We initiated a series of simulations based on our prior estimates of the size (N=300) and timing (30 generations ago) of the Ashkenazi bottleneck (Carmi et al., 2014; Palamara et al., 2012), and population-based estimates (Power et al., 2013) of reduced fecundity in schizophrenia (fecundity ratio ~0.5 for females and ~0.25 for males). We then generated 10,000 simulations for each of a series of variations on these parameters (Supplementary Table 5) in order to model the odds of a deleterious variant, present in a single individual at the time of the bottleneck, escaping extinction to persist in the present AJ population. In addition to the parameters noted above, simulations were performed as a function of penetrance for schizophrenia. As shown in Figure 2, between 30-50% of such variants escape extinction within the range of penetrance expected, given the genetic architecture of the disorder (Sullivan et al., 2012).

**Figure 2.**
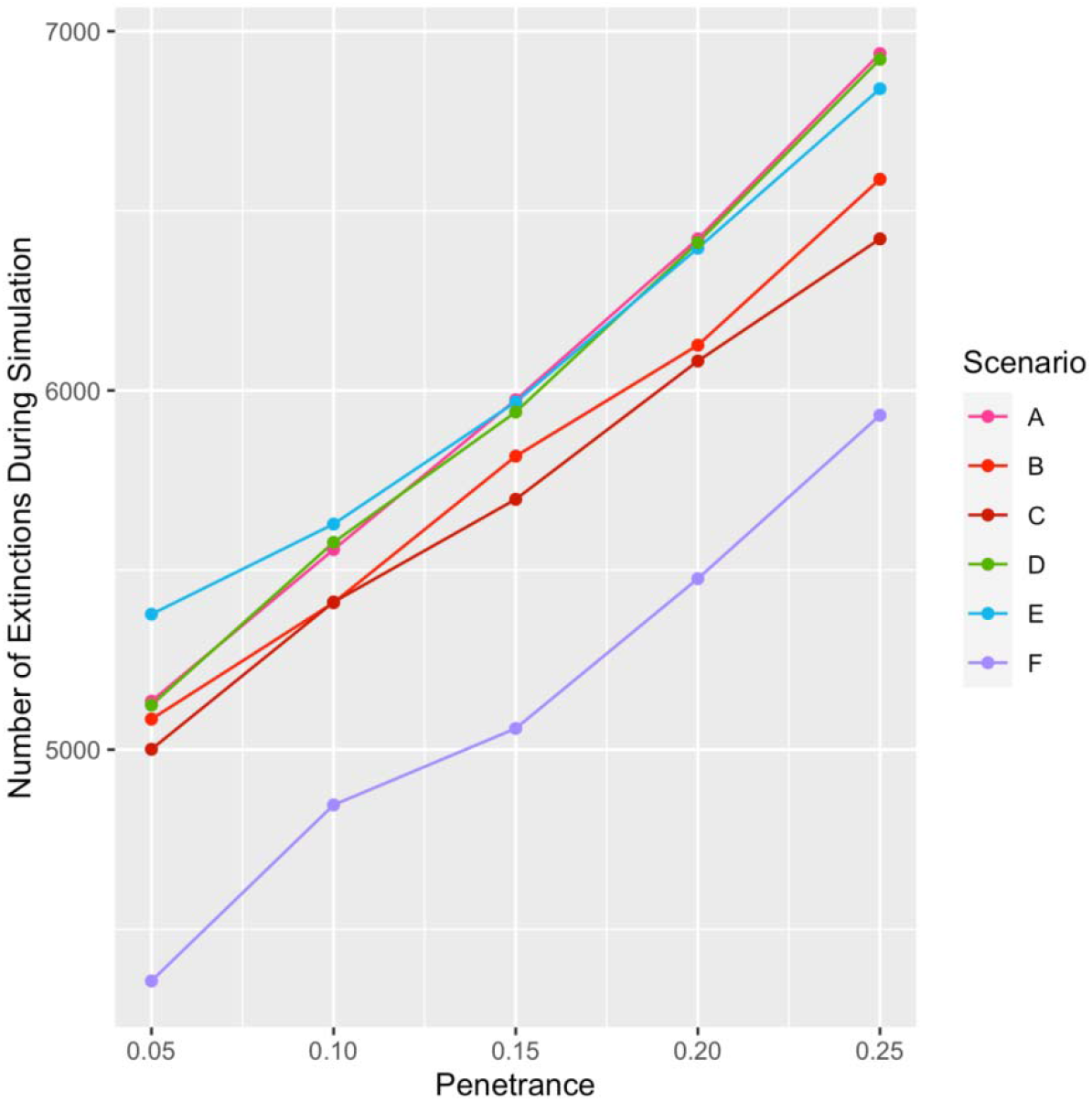
Between 30-50% of Damaging Variants Escape Extinction. Under a range of scenarios (see Supplementary Table 5) consistent with known population and disease parameters, as many as half of all damaging variants remain in a rapidly expanding founder population. Y-axis denotes number of extinctions out of 10,000 simulations for each condition.

Based on these results, and given rough estimates of the AJ population today (~10M) and the prevalence of schizophrenia (~1%), we could then estimate the total number of case and control carriers expected in the contemporary AJ population for each scenario. These calculations allowed us to determine the power of the present study to detect a given variant at exome-wide significance (Supplementary Figure 2), which generally ranged between 5% and 20%. However, we also calculated that a slightly larger study, of 5000 cases and 9000 controls, would have power of ~20-40% to detect any individual variant in the range of realistic penetrance (Supplementary Figure 3), and would therefore have >80% power to detect *at least* one variant, assuming there are at least 7 such variants circulating in the population (i.e., even if only 5% of our case-only list were true positives). Such an assumption is likely to be extremely conservative, given the estimated mutational target of 1000 genes or more (Nguyen et al., 2017; Purcell et al., 2014), the replication of the SCHEMA results in our dataset, the significant findings documented in Table 2, and the long list of variants at greater than doubleton frequency documented in Supplementary Table 5.

### MisLoF URVs Are Inversely Correlated with Common Variant Polygenic Risk Score in Cases

Finally, we tested the liability threshold model of schizophrenia, which suggests that genetic risk factors are largely additive; if true, then it would be expected that cases with a greater URV burden would require a lower burden of common risk variants, as indexed by GWAS-derived polygenic risk score (PRS). By contrast, no relationship between PRS and URV would be expected in controls. For each subject, PRS was calculated based on the large-scale schizophrenia GWAS reported by the Psychiatric Genomics Consortium (Schizophrenia Working Group of the Psychiatric Genomics Consortium, 2014), excluding our own Ashkenazi cohort. After controlling for sex and the first five principal components derived from the GWAS data, there was a significant inverse relationship between PRS and total number of MisLoF URVs in cases (β=-0.0217, s.e.=0.0096, p=0.024), but not in controls (β=-0.0094, s.e.=0.0129, p=0.465).

## Discussion

The present study demonstrates the enhanced power available to genetic studies performed in populations enriched for rare variants, consistent with recent work in schizophrenia (Gulsuner et al., 2020) and other phenotypes (Locke et al., 2019; Rivas et al., 2018; Selvan et al., 2020). We further reduced background heterogeneity by utilizing a strict filter against all variants reported in non-neuropsychiatric samples across the two largest publicly available sequencing datasets, gnomAD (Karczewski et al., 2020) and TOPMED (The NHLBI Trans-Omics for Precision Medicine (TOPMed) Whole Genome Sequencing Program., 2018). Thus, despite relatively modest sample sizes, the present study was able to replicate several previously identified schizophrenia-associated genes (*SETD1A, TRIO, XPO7)* (SCHEMA Consortium, 2020; Singh et al., 2016) and gene sets (synaptic, DBD-related, and constrained genes) (Genovese et al., 2016; Gonzalez-Mantilla et al., 2016; Gulsuner et al., 2020; Nguyen et al., 2017; Purcell et al., 2014), with these analyses serving as a positive control for our approach. Beyond these replications, we were also able to make several novel discoveries, as described below.

First, we identified several novel gene sets associated with schizophrenia. The strongest statistical signal was observed for cell adhesion processes, including cadherin family genes (Table 2b). Cadherins form calcium-dependent adherence junctions at the synapse and are involved in both neuronal migration and mature synaptic activity (Friedman et al., 2015). Surprisingly, cadherins have not received much attention in the schizophrenia genetics literature, despite the considerable recent focus on both calcium activity and synaptic proteins (Nanou & Catterall, 2018). While there are more than 100 different proteins in the cadherin superfamily (Friedman et al., 2015), it is noteworthy that three of the four FAT atypical cadherins, all in different chromosomal regions, appeared on our case-only list (as did their key interacting gene, *DCHS2)*. These genes are specifically involved in regulating microtubule polarity, thereby directing cellular migration in the developing nervous system (Avilés & Goodrich, 2017; Fulford & McNeill, 2020). Homozygous mutations in *FAT4* cause Van Maldergem syndrome, a recessive intellectual disability marked by periventricular neuronal heterotopia (Cappello et al., 2013), while mutations in *FAT1* have been observed in autism (Cukier et al., 2014).

Relatedly, a single missense variant in a protocadherin gene (*PCDHA3)* was observed at higher rate of recurrence (5 observations) in cases than any other ultra-rare (i.e., not in healthy individuals) variant in the published schizophrenia literature (although it should be noted that one splice acceptor variant in *SETD1A* appears six times in the SCHEMA database). *PCDHA3* is one of several protocadherins, clustered at a single locus on chromosome 5, which serve as a “molecular barcode” on the neuronal cell surface, guiding neurites away from forming synapses with other neurites from the same cell (Canzio & Maniatis, 2019). Altered expression of protocadherins (including *PCDHA3)* in schizophrenia has been implicated by a recent transcriptome-wide association study of both prefrontal cortex and hippocampus (Collado-Torres et al., 2019), and cortical interneurons derived from induced pluripotent stem cells (iPSCs) of patients with schizophrenia showed reduced *PCDHA3* expression compared to similarly derived interneurons from controls (Shao et al., 2019). The latter study further demonstrated that reduced protocadherin expression was associated with deficient synaptic arborization in both rodent and iPSC-derived human interneurons (but not glutamatergic neurons), and that these deficits could be reversed by treatment with an inhibitor of protein kinase C (Shao et al., 2019).

When our samples were combined with Ashkenazi patients from the SCHEMA database, the *PCDHA3* missense variant was observed in 0.3% of all Ashkenazi cases in the present study. While unusually high for a schizophrenia-associated URV, this carrier rate is low compared to the “4% rate observed for the most common *BRCA1* founder variant in Ashkenazi breast cancer cases (King et al., 2003), and the “15% rate of the *LRRK2* G2019S variant amongst Ashkenazi patients with Parkinson’s disease (Correia Guedes et al., 2010). These latter disorders have onset in late-life, and therefore susceptibility alleles for these diseases are not under the strong purifying selection affecting genes for schizophrenia (Pardiñas et al., 2018), a disorder which results in markedly reduced fecundity (Power et al., 2013). Nevertheless, our simulations demonstrated the limits of purifying selection in a founder population with a tight bottleneck. One-third to one-half of all damaging variants escape purifying selection, and these variants tend to become surprisingly frequent in the context of a rapidly expanding population, as described previously for the Finnish population (Wang et al., 2014). Consequently, ascertainment of additional samples from founder populations can be a highly cost-effective way of rapidly enhancing power of rare variant studies (Locke et al., 2019).

The overlap of our schizophrenia case-only gene list with gene sets derived from developmental brain disorders including autism and intellectual disability was notable, insofar as exome studies in these disorders have been more well-powered than schizophrenia studies to date (Myers et al., 2020). Consequently, the overlapping genes indicated in the first three rows of Table 2 (and especially the first row, which remained significant after permutation testing) have a strong prior probability of association, especially given prior evidence that rare single nucleotide variants (e.g., *SETD1A)* (Singh et al., 2016) and copy number variants (Kirov, 2015) tend to be shared across schizophrenia and other neurodevelopmental disorders. In the present study, case-only variants in the 11 genes overlapping the prior report of developmental brain disorders (DBD) (Gonzalez-Mantilla et al., 2016) (*ASXL3*, *BIRC6, DIP2A, DST, LAMA2, NSD1, PCDH15, SETBP1, SETD1A, TRIO, WDFY3)* were overwhelmingly (10:1 ratio) missense rather than loss of function; by contrast, the DBD list was generated from prior reports of loss of function variation exclusively. Thus, it is possible that our findings represent allelic series at these genes, in which more damaging variants are associated earlier-onset, more severe clinical phenotypes (Shohat et al., 2017). Similarly, we identified 5 cases (and no controls) with novel missense variants in *TSC2*, a gene in which mutations (primarily loss of function) are known to cause tuberous sclerosis (TS).

TS is an autosomal dominant disorder marked by hamartomas across multiple organs, potentially including the brain (Henske et al., 2016). Case reports of psychotic features in TS patients have proliferated for decades (Herkert et al., 1972); a recent survey of a large international cohort of TS patients identified psychosis in 11% of adults (de Vries et al., 2018). Since the affected cases in the present study were not noted in their medical report to have TS, our results suggest that schizophrenia can be the primary presenting feature of *TSC2* mutations.

In the last 15 years, genetic research in schizophrenia has given consistent support to the long-posited liability-threshold model (Gottesman & Shields, 1967; Kendler, 2015; McGue et al., 1983; Smeland et al., 2020), which states that manifestation of illness requires that the additive total of risk factors (including genetic and environmental) crosses an (unknown) threshold. While most schizophrenia research to date has focused on the total burden of common genetic variants, as captured by the polygenic risk score (Lee et al., 2012; The International Schizophrenia Consortium, 2009; van Rheenen et al., 2019), the model suggests that rare variants contribute in the same manner, albeit with much greater weighting (penetrance) (Richards et al., 2016). Supporting evidence has come from three very recent studies of schizophrenia patients carrying known, high-penetrance copy number variants (Bergen et al., 2019; Cleynen et al., 2020; Taniguchi et al., 2020). These studies all show that patients with highly penetrant CNVs have lower common-variant PRS compared to patients not carrying a known CNV, presumably because the CNV has already pushed them closer to the threshold for illness. In the present study, we have demonstrated, for the first time, an similar inverse correlation between common-variant PRS and rare variant burden indexed by missense and loss-of-function single nucleotide changes and small indels.

This study had several limitations, most notably that sample size was relatively small for a genetic association study; on the other hand, we demonstrated that the AJ population is enriched for rare variants and has substantially greater power than comparably-sized studies of outbred populations. Moreover, we were able to utilize external, well-powered datasets (i.e., SCHEMA and various studies of neurodevelopmental disorders) as validation/replication, and our gene set results for synaptic and constrained genes served as a positive control for our approach. Relatedly, although we utilized whole-genome sequencing to obtain our data, we restricted our analysis to the exome in order to make use of the largest possible set of samples both for purposes of these comparisons and for filtering of variants. Finally, we did not have neuropsychological testing data available for our cases, so we were unable to differentiate the relative contribution of URVs to cognitive deficits in our samples (Singh et al., 2017).

## Methods

### Subjects

Sequenced samples (n=1346) were derived from subjects described previously from multiple case-control cohorts summarized in Supplementary Table 7. All samples were self-reported to be Ashkenazi Jewish, and also verified as AJ by principal components analysis of previously collected SNP array data as described in our prior publications (Atzmon et al., 2010; Guha et al., 2012). Informed consent was obtained in accordance with institutional policies and the studies were approved by the corresponding institutional review boards.

Patients with schizophrenia were recruited from hospitalized inpatients at seven medical centers in Israel as described previously (Guha et al., 2013; Lencz et al., 2013). All diagnoses were assigned after direct interview using a structured clinical interview, a questionnaire with inclusion and exclusion criteria, and cross-references to medical records. The inclusion criteria specified that subjects had to be diagnosed with schizophrenia or schizoaffective disorder by the Diagnostic and Statistical Manual of Mental Disorders (DSM-IV). The exclusion criteria eliminated subjects diagnosed with at least one of the following disorders: psychotic disorder due to a general medical condition, substance-induced psychotic disorder, or any Cluster A (schizotypal, schizoid or paranoid) personality disorder. Controls were taken from several cohorts, primarily those screened for multiple forms of chronic illness (Lencz et al., 2013; Walter et al., 2011), but also including a small number of subjects ascertained for non-psychiatric disorders (Inflammatory Bowel Disease or Dystonia)(Kenny et al., 2012; Risch et al., 2007).

### Sequencing and variant calling pipeline

All samples were sequenced on the Illumina HiSeqX platform, using methods described previously (Lencz et al., 2018). Briefly, genomic DNA was isolated from whole blood and was quantified using PicoGreen, and integrity was assessed using the Fragment Analyzer (Advanced Analytical). Sequencing libraries were prepared using the Illumina TruSeq Nano DNA kit, with 100ng input, and pooled in equimolar amounts (8 samples/pool); a 2.5-3nM pooled library was loaded onto each lane of the patterned flow cell, and clustered on a cBot, generating ~375-400M pass filter 2×150bp reads per flow cell lane. For samples that did not meet 30x mean genome coverage post alignment, additional aliquots of the sequencing libraries were pooled in proportion to the amount of additional reads needed for re-sequencing.

Upon completion of sequencing runs, bcl files were demultiplexed and quality of sequencing data reviewed using SAV software (Illumina) and FastQC for deviations from expected values with respect to total number of reads, percent reads demultiplexed (>95%), percent clusters pass filter (>55%), base quality by lane and cycle, percent bases >Q30 for read 1 and read 2 (>75%), GC content, and percent N-content. FastQ files were aligned to hg19/GRCh37 using the Burrows-Wheeler Aligner (BWA-MEM v0.78)(Li & Durbin, 2009) and processed using the best-practices pipeline that includes marking of duplicate reads by the use of Picard tools (v1.83, http://picard.sourceforge.net), realignment around indels, and base recalibration via Genome Analysis Toolkit (GATK v3.5)(Van der Auwera et al., 2013). A total of 1,310 samples proceeded to the last steps of joint genotyping and VQSR variant filtering after removal of 10 samples (7 cases, 3 controls) with < 80% 20X read depth coverage of the genome, and removal of 26 duplicate samples (1 case and 25 controls) sequenced as part of QC procedures. All remaining samples were jointly genotyped to generate a multi-sample VCF. Variant Quality Score Recalibration (VQSR) was performed on the multi-sample VCF, and variants were annotated using VCFtools (Danecek et al., 2011). After the GATK pipeline, we also filtered the SNP and small INDEL on LCR regions (Li, 2014) and 1000G masked difficult regions (Auton et al., 2015), since these regions are enriched with calling errors that cannot be filtered effectively by the VQSR model, as we demonstrated previously (Lencz et al., 2018). Furthermore, we masked the genotypes with GQ < 20 as “./.” and filtered variants where <= 80% individuals could not be genotyped confidently. For purposes of downstream case-control analyses, we also removed 1 member of any pair of samples that were related at the first-cousin level or greater (n=27 controls).

### Defining ultra-rare variants (URVs) in TAGC samples

To maximize the power of existing reference population databases, we focused our primary analysis on the exome regions (as defined by gnomAD exome calling intervals (Karczewski et al., 2020)). We focused on URVs that were novel singletons in our TAGC cohort, filtering out all variants called in gnomAD (v2.1.1) non-neuro samples of any ethnicity, regardless of their call quality, and filtering out all variants called in TOPMed(The NHLBI Trans-Omics for Precision Medicine (TOPMed) Whole Genome Sequencing Program., 2018) freeze 5 release (with coordinates lifted over to hg19). We identified a small set of TAGC samples (n = 34; 21 cases / 13 controls) with an excessive number of exonic URVs (>=50), shown as outliers in the distribution (Supplementary Figure 1). Importantly, these outlier samples also demonstrated excess number of intergenic URVs and were not restricted to any sequencing batch. After filtering these outliers, we ended up with 1,249 samples for the down-stream analyses (Supplementary Table 7).

Primary analyses compared potentially functional (missense or loss of function) to putatively silent (synonymous or other) variants; these were defined as MisLoF and non-MisLoF, respectively. Exonic URVs were classified as loss of function, missense, synonymous, or other (generally intronic bases immediately flanking exons), based on their most damaging impact annotated for any transcript. Thus, non-MisLoF variants had no missense or loss of function annotation on any known transcript.

### Assigning novel URVs to genes and defining “case-only” and “control-only” genes

Each gene was characterized by the number of cases and the number of controls harboring a novel MisLoF or non-MisLoF variant within it. For each category of URV (MisLoF and non-MisLoF), we focused on genes in which only cases or only controls harbored a variant; genes in which both cases and controls were observed to have a given type of URV were excluded from subsequent analyses for that variant type. Of course, it was more likely that a gene would be identified as “case-only” rather than “control-only” for each type of URV due to the unequal numbers of cases relative to controls. Consequently, we controlled for the both effects by utilizing a re-sampling strategy, down-sampling the number of cases to match the number of controls, and iterated 10,000 times. For each iteration for each variant type, the following calculation was performed: (Case only genes - Control only genes))/(Case only genes + Control only genes).

### Replication of SCHEMA genes

The SCHEMA consortium lists 10 significant genes using strict exome-wide criteria (p<2.2×10^−6^) and 32 genes using false discovery rate <.05 (p<7.9×10^−5^)(SCHEMA Consortium, 2020). We used the hypergeometric test to statistically compare the overlap between case-only MisLoF genes in our AJ sample and these SCHEMA genes (restricting the comparison to the autosome). Note that we did not simply merge datasets due to differences in sequence acquisition, calling, and quality control procedures. To guard against potential confound by gene size, permutations were performed to match the size distribution of our case-only gene set. A permuted gene was randomly sampled from a window of ±25 in the order of coding sequence size for each gene in the set to be matched. A total of 10,000 iterations were performed, and the empirical p-value was defined by the proportion of the permuted gene set with overlap >= the case-only gene set. If no permutation was observed to meet this criteria, the p-value was reported as <1×10^−4^.

### Gene set analyses

We compared the AJ case-only and control-only MiSLoF genes to three categories of gene sets based on prior studies: 1. *De novo* mutation genes implicated across multiple developmental brain disorders (DBD)(Gonzalez-Mantilla et al., 2016), a large scale autism spectrum disorder(ASD) exome study(Satterstrom et al., 2020), and the integration of the ASD set with a large-scale study of ASD, developmental disorder(DD), and intellectual disability(ID) exome sequencing studies (Coe et al., 2019); 2. Genes known to encode proteins of the synapse aggregated in SynaptomeDB (Pirooznia et al., 2012), and genes regulated by known neuronal RNA-binding proteins, including CELF4 (Wagnon et al., 2012), FMRP (Darnell et al., 2011), RBFOX2 and RBFOX1/RBFOX3 (Weyn-Vanhentenryck et al., 2014); 3. Genes constrained by missense (Samocha et al., 2014) and LoF variants (Karczewski et al., 2020). Since X and Y chromosomes of AJ samples are not included, we adjusted the number of genes in each set and total protein-coding genes (n=19,780; http://hgdownload.cse.ucsc.edu/goldenPath/hg19/database) to autosomal-only to calculate the p-values using hypergeometric tests. We further compared the relative enrichment between AJ case-only and control-only MisLoF genes in the prior gene sets using the chi-squared test based on the 2×2 table of overlap and non-overlap for case-only and control-only. As above, we also performed permutation testing to control for effects of gene size.

### Modeling the effects of purifying selection

We simulated the spread of a single schizophrenia-causing variant in a founder population under a number of empirical conditions. The initial conditions of the simulation assume a single variant carrier in a fixed size population at the time of bottleneck, which we denote as *N_B_*. From there, we modeled the growth of the number of carriers and total population size over a set number of *G* generations until a maximum population size *N_max_* is achieved. The growth rate *R* is computed as *R = e(ln(N_max_/N_B_)/G)*. We generated the number of offspring per individual within a generation using a Poisson branching process, in which the lambda parameter is a function of growth rate *R*, sex, and case/control status. A healthy individual will have a lambda equal to *R*, while an individual with schizophrenia will have reduced relative fecundity, differing by sex. Within our simulated population, we track the number of variant carriers while varying the population bottleneck size *N_B_*, number of generations *G*, relative fecundity of schizophrenia cases, and the disease penetrance of the variant. We computed 10,000 simulations for each scenario, calculating the proportion of simulations in which the number of variant carriers goes to zero (extinction) within *G* generations, and the proportion of simulations in which a variant escapes extinction. We then use the number of variant carriers remaining after *G* generations in the population to calculate the power of Fisher’s exact test for detecting the variant effect in a study cohort size of a given size.

### Common variant polygenic risk score (PRS)

A common variant PRS for schizophrenia was calculated for each subject based on summary statistics from the large-scale schizophrenia GWAS reported by the Psychiatric Genomics Consortium (Schizophrenia Working Group of the Psychiatric Genomics Consortium, 2014), excluding our own Ashkenazi cohort. Regression analysis examining the relationship between PRS and total number of MisLoF URVs, controlling for sex and the first five genetic principal components derived from the GWAS data, was performed separately for cases and controls.

## Data Availability

Variant files will be made available through the European Genome-Phenome Archive (EGA) upon peer-reviewed publication, consistent with our prior work (https://ega-archive.org/datasets/EGAD00001000781).

## Acknowledgements

The authors are extremely grateful to Soren Germer, Ph.D. and his team at the New York Genome Center for performing the Illumina sequencing. We acknowledge financial support from the Human Frontier Science Program (SC); NIH research grants AG042188 (GA), DK62429, DK062422, DK092235 (JHC), NS050487, NS060113 (LNC), AG021654, AG027734 (NB), MH089964, MH095458, MH084098 (TL), and CA121852 (computational infrastructure, IPe’er); NSF research grants 08929882 and 0845677 (IPe’er); Rachel and Lewis Rudin Foundation (HE); Northwell Health Foundation (TL); Brain & Behavior Foundation (TL); US-Israel Binational Science Foundation (TL, AD); LUNGevity Foundation (ZHG); New York Crohn’s Disease Foundation (IPeter); Edwin & Caroline Levy and Joseph & Carol Reich (SB); the Parkinson’s Disease Foundation (LNC); the Sharon Levine Corzine Cancer Research Fund (KO); and the Andrew Sabin Family Research Fund (KO).

## Author Contributions

TL and IP led the analysis, and TL led the writing of the manuscript. JY and RRK conducted the primary analyses, with assistance from ML and SC. TL led the funding of the study. TL, AD, GA, DB, NB, and LNC provided samples and conducted lab work. TL, IP, NB, SB, AD, JHC, LNC, ZHG, VJ, RK, SL, KO, HO, LJO, IP, AMK, and GA initiated and designed the study, and provided funding.

Competing financial interests:

The authors declare no competing financial interests.

**Supplementary Table 2.**

|  | **Genes in**<br>**Gene_set** | **Case-only**<br>**overlap** | **Control-only**<br>**overlap** | **Permutation**<br>**P(Case)** | **Permutation**<br>**P(control)** |
| --- | --- | --- | --- | --- | --- |
| DBD(Gonzalez-Mantilla et al., 2016) | 214 | 11 | 1 | 0.0034 | 0.9534 |
| ASD(Satterstrom et al., 2020) | 102 | 4 | 0 | 0.1404 | 1 |
| ASD_ID_DD<br>(Coe et al., 2019; Satterstrom et al., 2020) | 274 | 8 | 2 | 0.1429 | 0.7887 |
| synaptome(Pirooznia et al., 2012) | 1828 | 26 | 15 | $3.00 \times 10^{-4}$ | 0.2102 |
| CELF4(Wagnon et al., 2012) | 2504 | 41 | 23 | $<1 \times 10^{-4}$ | 0.1723 |
| FMRP(Darnell et al., 2011) | 1210 | 35 | 19 | $3.00 \times 10^{-4}$ | 0.2719 |
| RBFOX2(Weyn-Vanhentenryck et al., 2014) | 2911 | 48 | 27 | $<1 \times 10^{-4}$ | 0.2131 |
| RBFOX13(Weyn-Vanhentenryck et al., 2014) | 3255 | 50 | 27 | $<1 \times 10^{-4}$ | 0.325 |
| Missense-constrained(Samocha et al., 2014) | 961 | 23 | 8 | 0.0092 | 0.8221 |
| LoF-constrained (Karczewski et al., 2020) | 3264 | 62 | 35 | $<1 \times 10^{-4}$ | 0.1974 |

**Supplementary Table 3.**

| **GO cellular component** | **total genes** | **overlap** | **expected** | **raw p** | **FDR** |
| --- | --- | --- | --- | --- | --- |
| neuron projection (GO:0043005) | 1381 | 23 | 9.27 | $5.00 \times 10^{-5}$ | $2.51 \times 10^{-2}$ |
| synapse (GO:0045202) | 1373 | 22 | 9.22 | $1.30 \times 10^{-4}$ | $2.60 \times 10^{-2}$ |
| cell junction (GO:0030054) | 2103 | 31 | 14.12 | $2.19 \times 10^{-5}$ | $2.19 \times 10^{-2}$ |
| plasma membrane bounded cell projection (GO:0120025) | 2208 | 31 | 14.83 | $5.55 \times 10^{-5}$ | $2.23 \times 10^{-2}$ |
| cell projection (GO:0042995) | 2296 | 32 | 15.42 | $4.70 \times 10^{-5}$ | $3.14 \times 10^{-2}$ |
| cytoskeleton (GO:0005856) | 2278 | 31 | 15.3 | $9.91 \times 10^{-5}$ | $2.21 \times 10^{-2}$ |
| vesicle (GO:0031982) | 3886 | 45 | 26.09 | $9.19 \times 10^{-5}$ | $2.30 \times 10^{-2}$ |
| organelle (GO:0043226) | 13720 | 117 | 92.12 | $2.14 \times 10^{-6}$ | $4.28 \times 10^{-3}$ |
| cellular anatomical entity (GO:0110165) | 18633 | 137 | 125.11 | $1.32 \times 10^{-4}$ | $2.41 \times 10^{-2}$ |
| cellular_component (GO:0005575) | 18824 | 138 | 126.39 | $7.80 \times 10^{-5}$ | $2.61 \times 10^{-2}$ |
| Unclassified (UNCLASSIFIED) | 2027 | 2 | 13.61 | $7.80 \times 10^{-5}$ | $2.23 \times 10^{-2}$ |

**Supplementary Table 4.**

| **Cellular Component** | **Gene count** | **p-value** | **q-value** |
| --- | --- | --- | --- |
| Synapse | 18 | $6.19 \times 10^{-4}$ | $2.25 \times 10^{-3}$ |
| presynaptic active zone | 5 | $7.49 \times 10^{-4}$ | $2.25 \times 10^{-3}$ |
| Presynapse | 10 | $4.72 \times 10^{-3}$ | $9.45 \times 10^{-3}$ |
| integral component of postsynaptic density membrane | 3 | $2.84 \times 10^{-2}$ | $4.27 \times 10^{-2}$ |
| **Biological Process** |  |  |  |
| Synapse | 18 | $6.19 \times 10^{-4}$ | $2.25 \times 10^{-3}$ |
| presynaptic active zone | 5 | $7.49 \times 10^{-4}$ | $2.25 \times 10^{-3}$ |
| Presynapse | 10 | $4.72 \times 10^{-3}$ | $9.45 \times 10^{-3}$ |
| integral component of postsynaptic density membrane | 3 | 0.0284 | 0.0427 |
| Postsynapse | 8 | 0.0935 | 0.1122 |

**Supplementary Table 5.**

| **N Cases** | **Gene** | **Chr** | **Pos** | **Ref** | **Alt** | **Impact** |
| --- | --- | --- | --- | --- | --- | --- |
| 5 | *PCDHA3* | 5 | 140182458 | A | G | missense |
| 4 | *ACBD6* | 1 | 180382593 | T | G | missense |
| 4 | *IGF1R* | 15 | 99251324 | C | T | missense |
| 3 | *ATAD3C* | 1 | 1389777 | CA | C | frameshift |
| 3 | *ACOX3* | 4 | 8412048 | G | A | missense |
| 3 | *FIGNL1* | 7 | 50513662 | G | A | missense |
| 3 | *CEP104* | 1 | 3740011 | T | C | missense |
| 3 | *UBR4* | 1 | 19426131 | A | G | missense |
| 3 | *KLHL30* | 2 | 239049577 | T | C | missense |
| 3 | *TBX18* | 6 | 85457687 | G | C | missense |
| 3 | *CACNA2D1* | 7 | 81599254 | A | C | missense |
| 3 | *TENM4* | 11 | 78369465 | A | T | missense |
| 3 | *TNFRSF1A* | 12 | 6440026 | C | A | missense |
| 3 | *BCAT1* | 12 | 25002856 | T | C | missense |
| 3 | *LCP1* | 13 | 46722531 | C | T | missense |
| 3 | *PCSK2* | 20 | 17240934 | A | T | missense |
| 3 | *PTK6* | 20 | 62164958 | A | G | missense |

**Supplementary Table 6.**
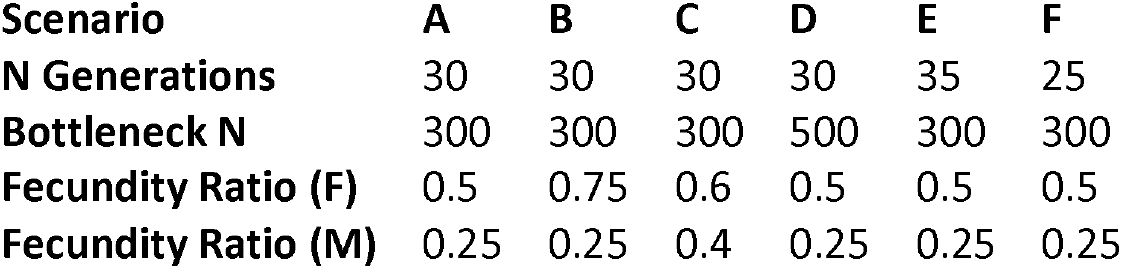

| **Scenario** | **A** | **B** | **C** | **D** | **E** | **F** |
| --- | --- | --- | --- | --- | --- | --- |
| **N Generations** | 30 | 30 | 30 | 30 | 35 | 25 |
| **Bottleneck N** | 300 | 300 | 300 | 500 | 300 | 300 |
| **Fecundity Ratio (F)** | 0.5 | 0.75 | 0.6 | 0.5 | 0.5 | 0.5 |
| **Fecundity Ratio (M)** | 0.25 | 0.25 | 0.4 | 0.25 | 0.25 | 0.25 |

**Supplementary Table 7.**

| Investigator(s) | All sequenced | Passed for joint-calling | After removing related | Included in final analysis | Female | Male | Phenotypes | Paper describing samples (PMID) |
| --- | --- | --- | --- | --- | --- | --- | --- | --- |
| Todd Lencz, Ariel Darvasi | 815 | 807 | 807 | 786 | 283 | 503 | Schizophrenia cases | 24253340 |
| Todd Lencz, Ariel Darvasi | 132 | 132 | 131 | 130 | 83 | 47 | controls | 24253340 |
| Gil Atzmon | 331 | 304 | 301 | 298 | 178 | 120 | controls | 21782286 |
| Judy Cho, Inga Peter | 34 | 33 | 23 | 16 | 6 | 10 | IBD cases | 22412388 |
| Laurie Ozelius | 34 | 34 | 21 | 19 | 10 | 9 | Dystonia cases | 17503336 |
| **TOTAL** | **1346** | **1310** | **1283** | **1249** | **560** | **689** |  |  |

**Supplementary Figure 1.**
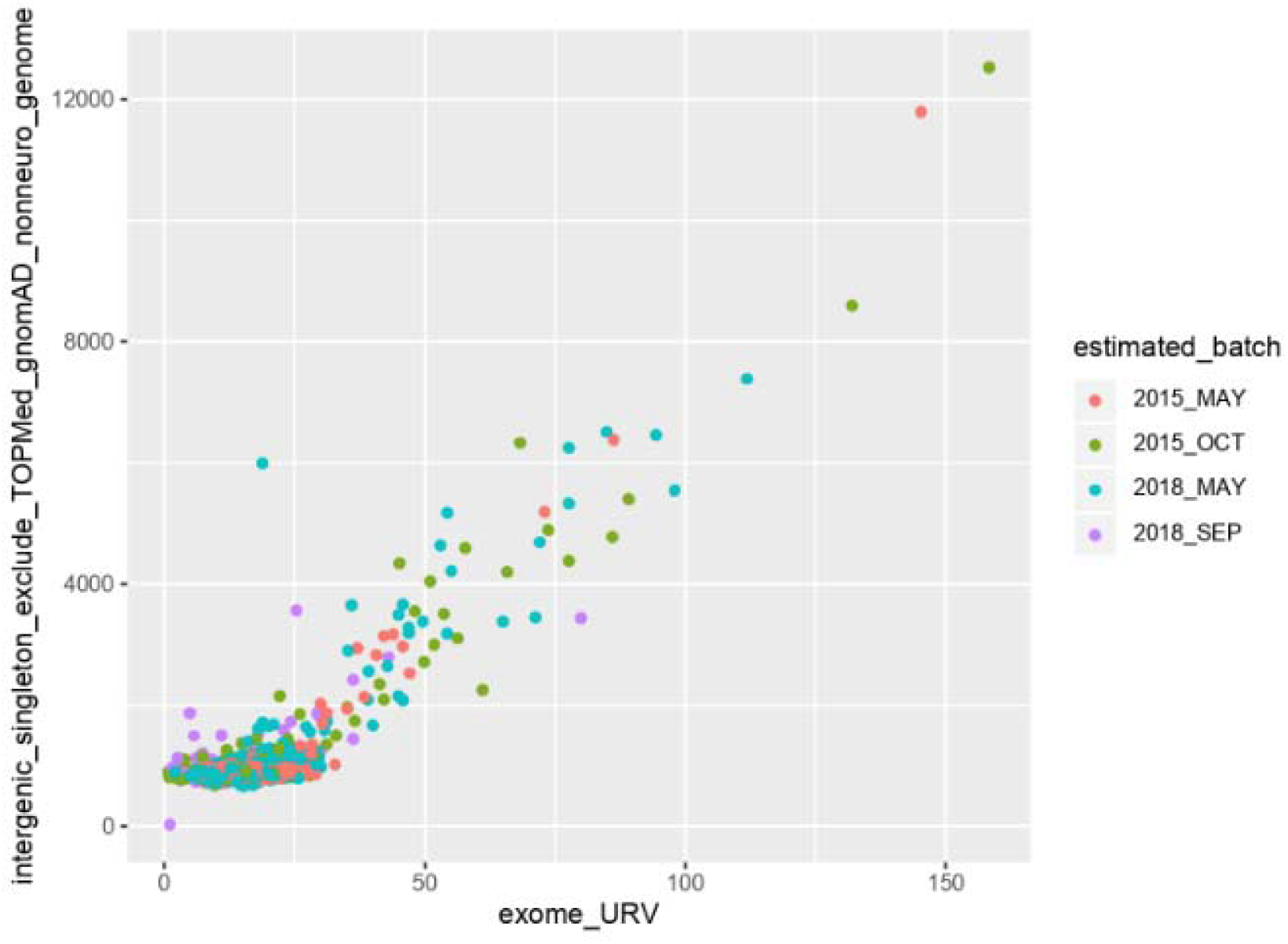

**Supplementary Figure 2.**
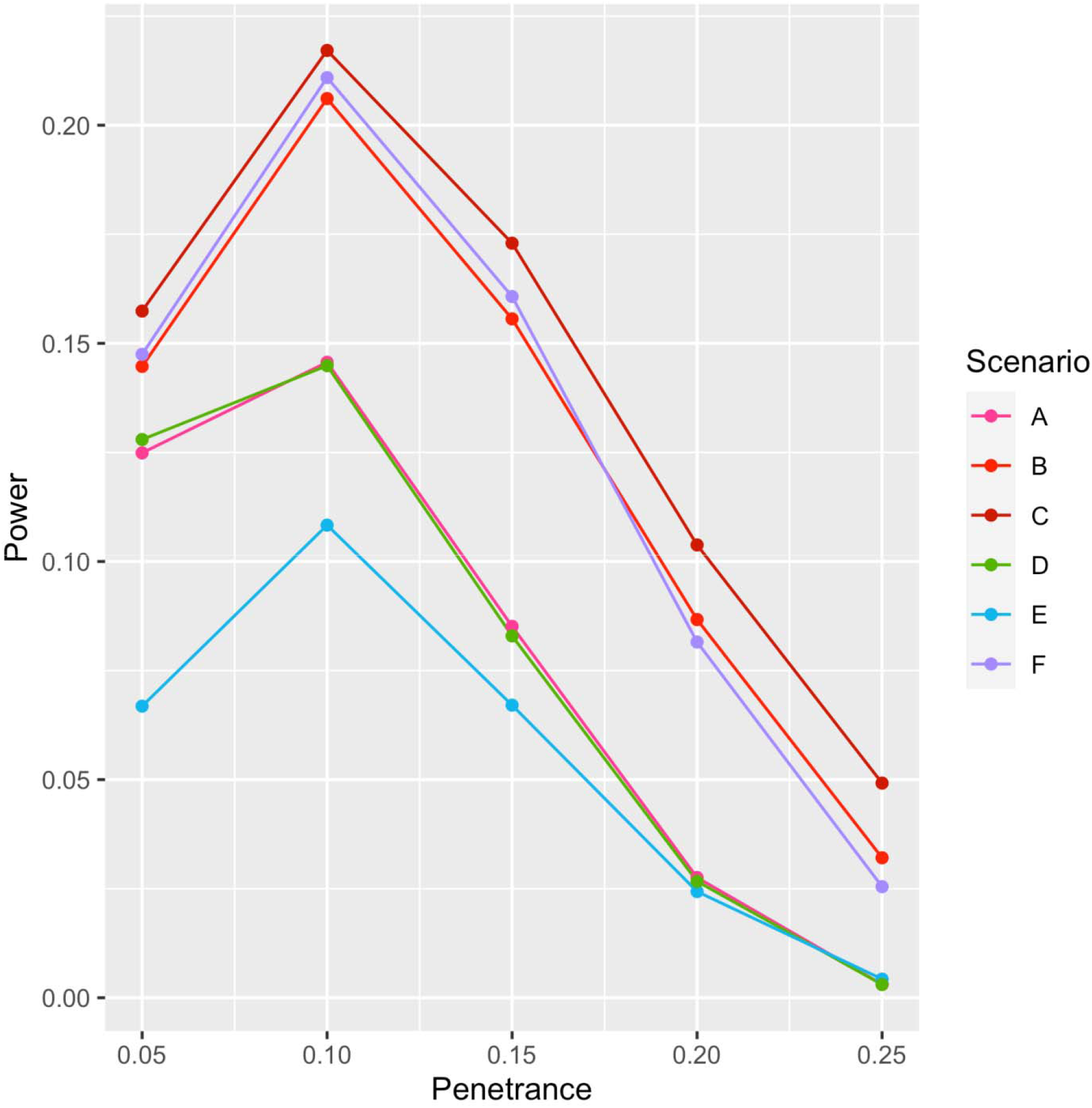

**Supplementary Figure 3.**
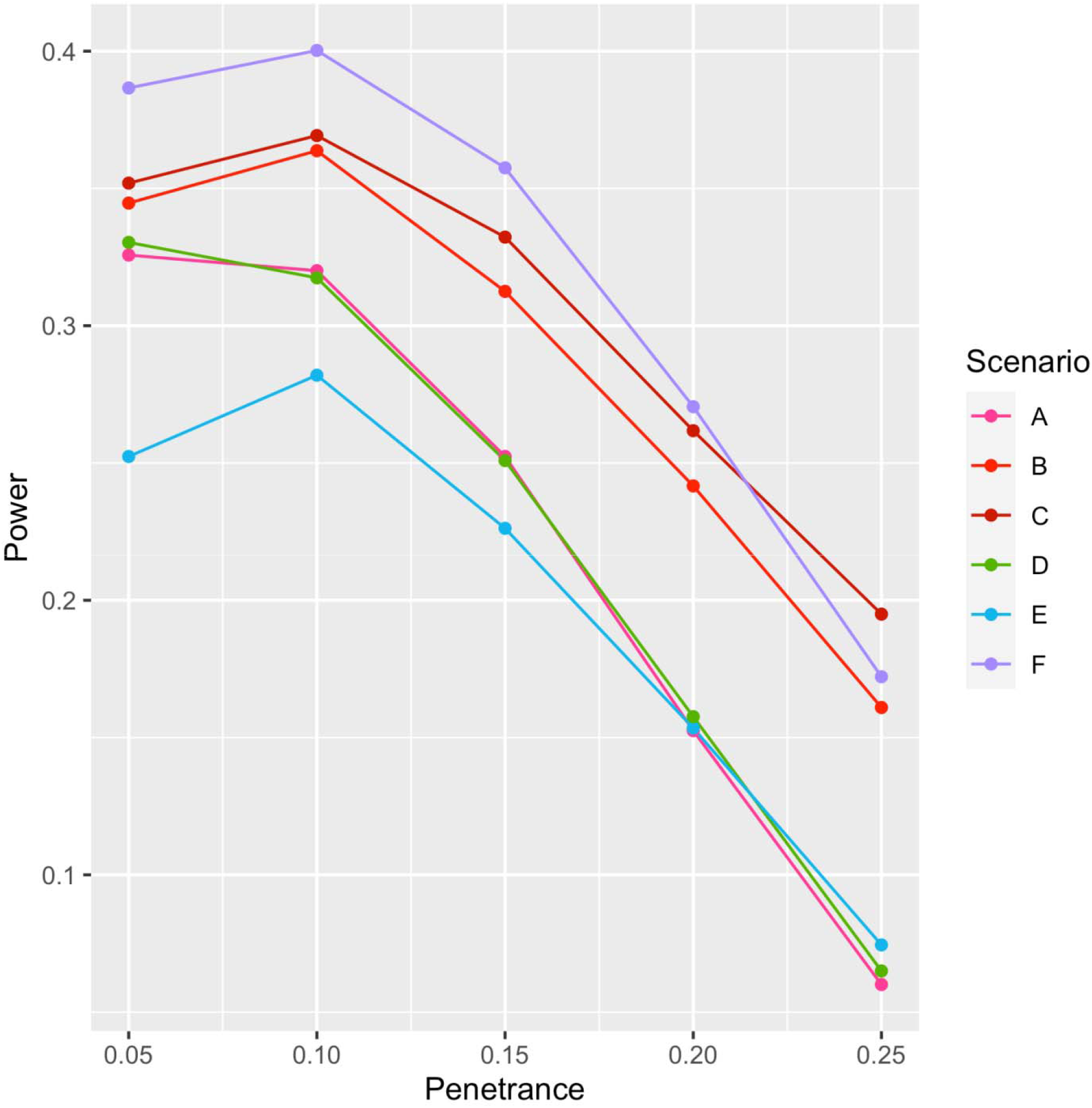

